# A scalable model for Parkinson’s disease genetic research in underrepresented populations: lessons from Kyrgyzstan

**DOI:** 10.64898/2026.09.08.26362411

**Authors:** Cholpon Shambetova, Eliza Zhunusova, Aizhamal Ismailova, Maria Teresa Periñan, Suium Kiyalbekova, Carolin Gabbert, Shen-Yang Lim, Gulmira Djumalieva, Aizhamal Iliazova, Nuraiym Beishembieva, Aruuke Kubanychbekova, Yuliia Kanana, Ekaterina Peredereeva, Adilet Metinov, Aleksandr Sorokin, Aisuikum Abdumalikova, Shohista Bazarbaeva, Gulnaz Pakhridinova, Batma Sattarova, Aiza Upaeva, Gulmira Zhamilova, Eliza Zhumadylova, Elmira Mamytova, Mirbek Baisalov, Altynai Baitelieva, Jyldyz Imanalieva, Aida Bolotalieva, Anara Toktomametova, Nargiza Atambekova, Sabina Baltabaeva, Aziza Dalbaeva, Zhumagul Osmonova, Bermet Nurbekova, Aliia Mukhanova, Irina Tsopova, Teresa Kleinz, Irina Sverdlova, Lara Mariah Lange, Joanne Trinh, Cornelis Blauwendraat, Andrew Singleton, Thomas F. Münte, Alastair J Noyce, Indira Kudaibergenova, Christine Klein, Global Parkinson’s Genetics Program (GP2)

## Abstract

**Background:** Scalable models for Parkinson’s disease (PD) genetics in underrepresented populations are needed, as most studies have focused on individuals of European-ancestry, limiting generalisability across ancestral and environmental contexts. Kyrgyzstan has been largely absent from global PD research. Using Kyrgyzstan as a case study, we aimed to develop and evaluate a scalable PD genetics model integrating research and local capacity building.

**Methods:** We implemented a stepwise capacity-building framework within a multi-site prospective PD genetics cohort that included participants with PD and controls, with longitudinal follow-up. The framework integrated community engagement, workforce development, specialist clinical services, data governance, clinical phenotyping, biobanking, and genetic data generation. Participants were recruited through a university-affiliated movement disorders clinic and mobile outreach clinics, using a secure electronic data-capture system and Global Parkinson’s Genetics Program (GP2)-aligned standards. Feasibility endpoints included recruitment, participation, assessment completeness, and genetic quality-control metrics.

**Findings:** Within 20 months, specialist clinical services and a PD registry with biobanking and secure data infrastructure were established, and 1240 participants were enrolled, representing multiple Central Asian ethnicities, including Kyrgyz, Uzbek, Uyghur, Tatar, Kalmyk, and Dungan, and the country’s rural– urban distribution. High participation coincided with community engagement and recruitment through mobile clinics and community outreach.

**Interpretation:** Embedding capacity building within active research enabled rapid development of neurogenetic and specialist clinical infrastructure in a resource-limited setting. This programme provides a potentially transferable model for equitable inclusion of underrepresented populations in PD genetics and related neurogenetic research in low- and middle-income countries.

**Funding:** This project was supported by the Global Parkinson’s Genetics Program (GP2; https://gp2.org). GP2 is funded by the Aligning Science Across Parkinson’s (ASAP) initiative and implemented by The Michael J. Fox Foundation for Parkinson’s Research (MJFF)

**Research in context:** *Evidence before this study:* We searched PubMed/MEDLINE and Scopus from 1995 to July 2026 for articles and reviews using combinations of terms related to Parkinson’s disease (PD), genetics, genomics, underrepresented populations (URP), ancestry, diversity, implementation, research infrastructure, capacity building, governance, and consortium development. Additional relevant publications were identified by reviewing the reference lists of pertinent articles. Eligible papers were categorized by study design, focus on implementation, implementation domains, and relevance to this research (Supplementary Table S1). Most available work comprises genome wide association studies, genetic discovery research, and reviews that address the lack of diversity in PD genetics. Several studies documented persistent ancestry disparities and underrepresentation of URP and called for broader inclusion, while regional reviews summarised evidence from Africa, Latin America, and other underserved regions. Few publications examined implementation issues such as international partnerships, governance, workforce training, consortium structures, or data sharing systems. Initiatives including the Global Parkinson’s Genetics Program (GP2), the International Parkinson Disease Genomics Consortium (IPDGC), the African and Arabian Parkinson’s Disease Genetics Consortium (AA PD GC), and the Michael J. Fox Foundation Global Genetic Parkinson’s Disease Project demonstrated that coordinated consortia can be built successfully. However, there is little published guidance on how to design and run integrated PD genomic programmes for URP, particularly in low resource settings or in Central Asia.

*Added value of this study:* This study describes the prospective development and assessment of an integrated genomic programme for PD embedded within routine neurological care in Kyrgyzstan, a Central Asian population previously unrepresented in PD genetics research. The project established a nationwide cohort and biobank, together with governance frameworks, workforce training, standardised clinical phenotyping, biospecimen collection, data management, community engagement, and routes for international data sharing. By analysing data collected during the setup phase, we identify operational obstacles and propose practical solutions to support sustainable integration of genomic research into clinical practice. Rather than focusing only on genetic discoveries, we outline an implementation model that could be adapted for similar URP.

*Implications of all the available evidence:* Current literature highlights the scientific importance of increasing ancestral diversity in PD genetics but offers limited direction on how to build and sustain such research in underrepresented regions. Our findings suggest that successful implementation depends on coordinated development of clinical services, governance, workforce capacity, biobanking, data infrastructure, community engagement, and international collaboration alongside genomic research. The model presented here may help guide future efforts to improve equitable participation in PD genetic studies and to strengthen research capacity in other resource-limited and underserved settings.

## Introduction

Despite rapid advances in human genomics, most genomic studies still disproportionately represent populations of European ancestry, limiting both scientific discovery and equitable translation of genomic medicine into clinical practice and public health.^1^ This gap is evident in Parkinson’s disease (PD), where recent genomic efforts have demonstrated that, until very recently, 91% of reported monogenic PD cases were derived from individuals of Northern European ancestry.^2^ Greater inclusion of ethnically and geographically diverse populations in genomic research is essential not only for equitable participation but also for improving understanding of disease biology and ensuring that genomic discoveries are applicable across diverse populations. Expanding ancestry representation enables the systematic study of genetic contributions to disease susceptibility, progression, and resilience across diverse populations, uncovering pathogenic mechanisms, modifier effects, and protective variants that remain undetectable in European population-centered datasets. In PD, where genetic findings increasingly inform risk stratification, genetic counselling, target prioritisation for drug development, and clinical trial enrolment, limited diversity risks inequitable implementation of precision medicine approaches across health systems.^3^

The study of PD and parkinsonism in underrepresented populations (URP) already provides multiple instructive examples of this principle. X-linked dystonia-parkinsonism (XDP), a condition exclusively present in individuals of Filipino descent tracing their origin to the Island of Panay, has served as a model disease for genetic modifier detection^4–6^ with implications for other, much more common diseases, such as Huntington’s disease or SCA27B ataxia^7^. Other examples include marked diversity in the distribution of pathogenic and risk variants in the *LRRK2* and *GBA1* genes across the globe, including the recent discovery within the Global Parkinson’s Genetics Program (GP2)^8^ of a common, ancestry-specific *GBA1* risk variant in individuals of African and African-admixed ancestry,^9^ which has led to new causal and pathophysiological insights^10^. Crucially, each of these advances depended on sustained local clinical and research capacity supported by long-term international partnerships, underscoring the limitations of short-term, extractive research models and highlighting that inclusive discovery requires sustained investment in locally led research infrastructure.

Central Asia remains one of the least represented regions in PD and genomic research despite its large population, genetic diversity, and distinct environmental exposures. Kyrgyzstan, a lower-middle-income country (LMIC) in Central Asia with a population of approximately 7·3 million, a predominantly rural demographic profile, and a multiethnic population (Figure 1), exemplifies both the challenges and opportunities of expanding PD genomics to underrepresented settings. As in many LMICs, progress in PD genomic research has been constrained by limited specialist capacity, gaps in expertise and research infrastructure, and low prioritization of neurological non-communicable diseases. In contrast, donor-supported genomic programmes for infectious diseases, have demonstrated that comparable capacity can be developed when aligned with national health priorities. Recent policy developments, including strengthened personal data legislation and adherence to the Nagoya Protocol, further support ethical international genomic research partnerships. Establishing sustainable PD genomic research in underserved settings requires coordinated development of clinical services, workforce capacity, governance, laboratory infrastructure, and international partnerships.

**Figure 1.**
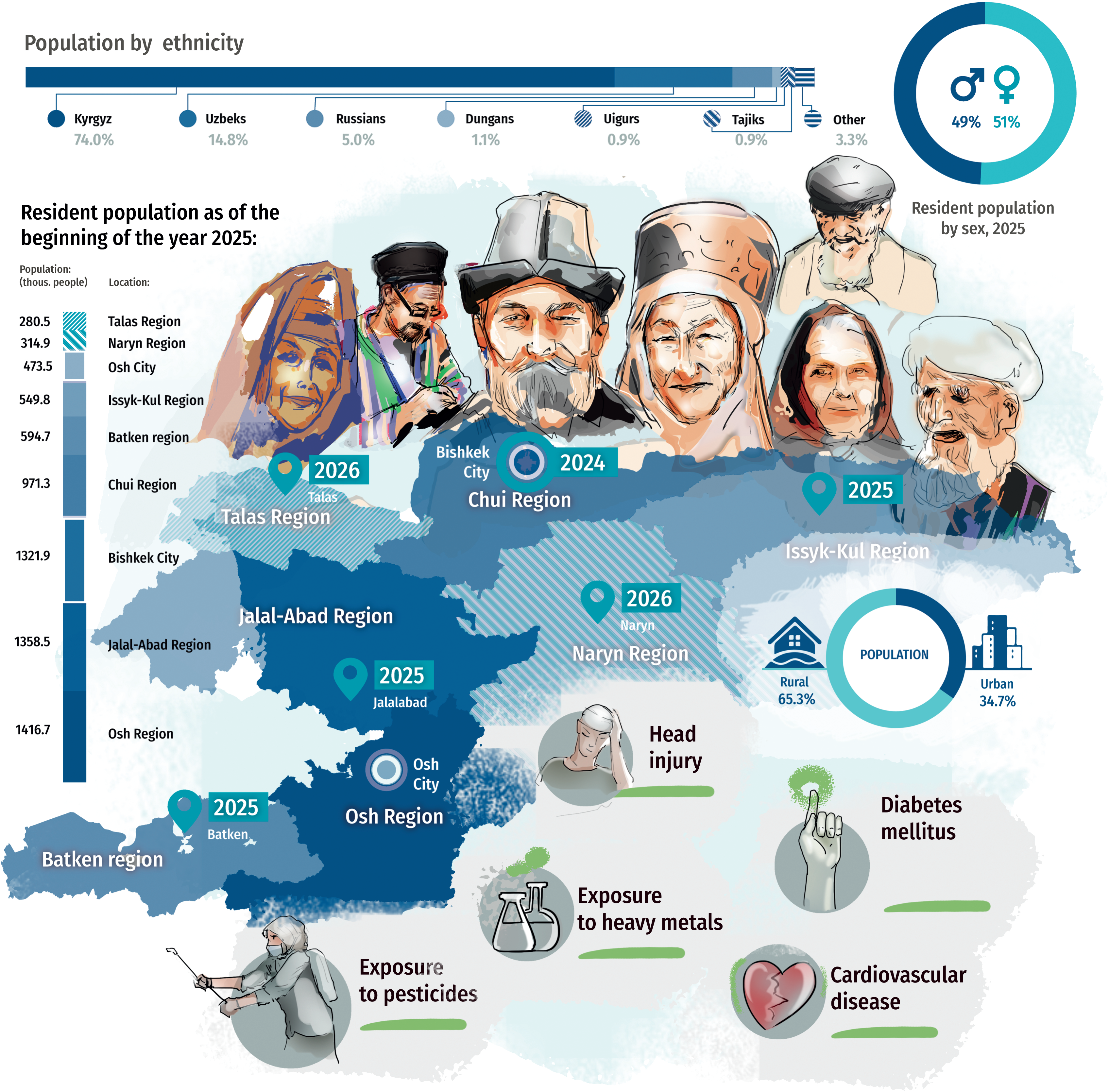
Overview of populations and ethnicities in Kyrgyzstan. The figure shows the geographic regions of Kyrgyzstan, the locations of main clinical sites, and outreach regions served by mobile visits (with year of establishment), alongside the country’s major ethnic groups. Selected lifestyle-related Parkinson’s disease (PD) risk factors are indicated for contextual reference. Footnote: Regions are color-coded according to population density. Ethnic groups representing less than 0·89% of the national population are not shown individually and are aggregated for clarity. Ethnic composition is based on national census data and reflects self-identified ethnicity. Geographic boundaries are schematic and intended for contextual illustration only.

At the outset of this work, Kyrgyzstan had no established infrastructure for PD genomic research. Over the subsequent two years, we developed an integrated PD genomic research programme that combined capacity building through GP2 (Figure 2; Supplementary Figure 1). Here, we describe its development, implementation, early evaluation, and initial cohort characteristics, and assess its potential to inform similar initiatives in other URP.

**Figure 2.**
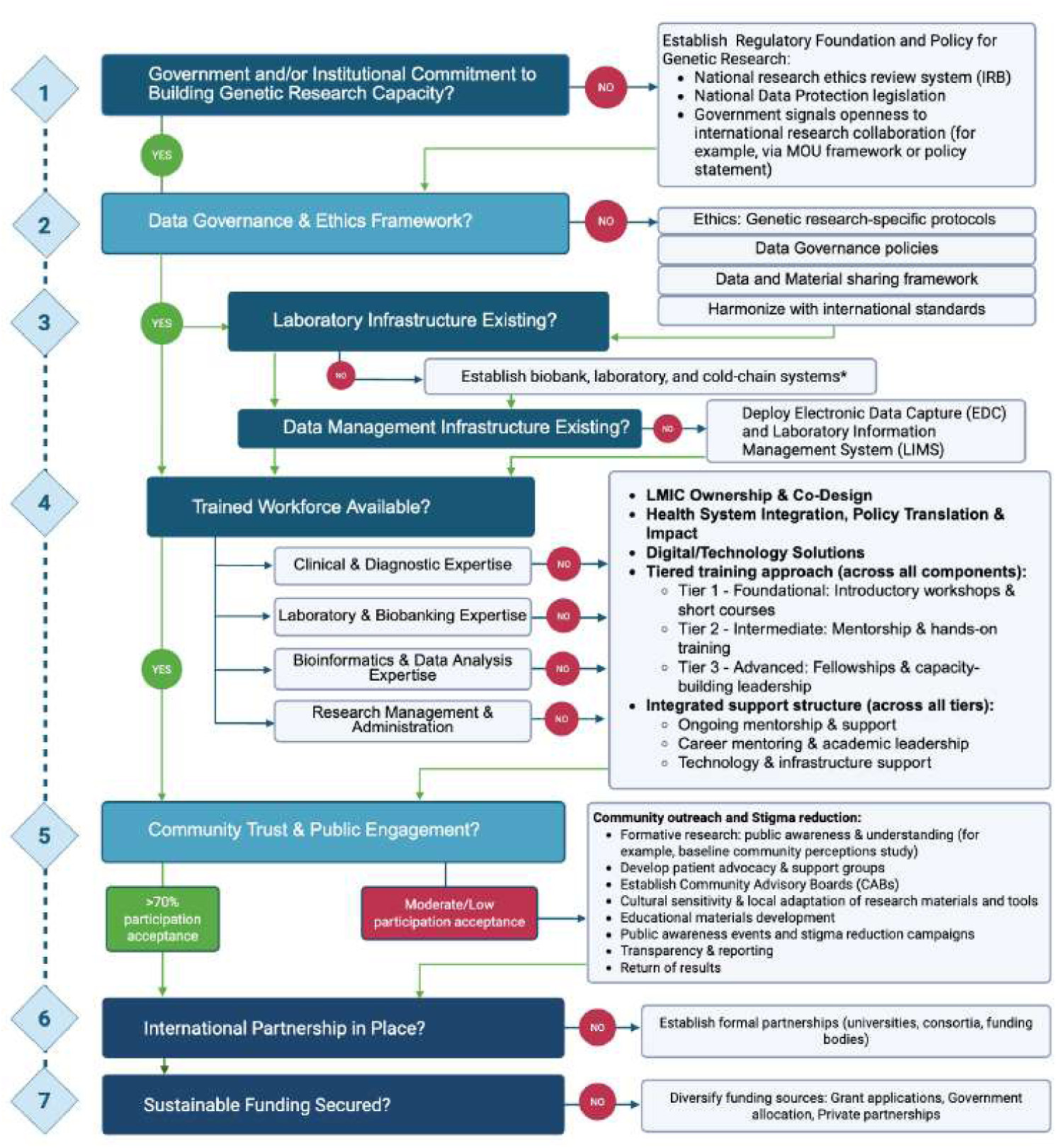
Conceptual framework for staged development of genetic research capacity in under-represented settings. The framework outlines key decision points and enabling components across governance, ethics, infrastructure, workforce development, community engagement, international partnership, and sustainable funding. Although presented in a staged format for clarity, the framework is intended to guide implementation rather than prescribe a fixed sequence; in practice, components may be implemented in parallel or iteratively depending on local context. While informed by the Kyrgyzstan implementation, the framework is designed as a generalisable scaffold adaptable to other low- and middle-income and under-represented settings. Created in https://BioRender.com

## Methods

### Study design and capacity-building framework

The study was conducted as part of the GP2, led by the I.K. Akhunbaev Kyrgyz State Medical Academy (KSMA) in partnership with the University of Luebeck (UzL) and in collaboration with the Ministry of Health (MoH) of the Kyrgyz Republic. We developed a staged framework for establishing genetic research capacity in URP (Figure 2), informed by relevant literature on global neuroscience capacity strengthening and implementation science in LMICs^11–13^. Components relevant to scalability and transferability are summarised in Supplementary Table S2. The framework comprised seven domains: (1) institutional and government alignment; (2) ethics, data, and material governance; (3) laboratory, biobanking, and data infrastructure; (4) workforce training; (5) community engagement; (6) international partnerships; and (7) long-term sustainability. Domains were implemented in parallel or iteratively. Progress was assessed against implementation deliverables, including regulatory approvals, infrastructure readiness, trained personnel, and data governance.

The primary research component comprised a multisite case–control study of patients with PD and Parkinson-plus syndromes (PPS) and healthy controls, with longitudinal follow-up of patients with parkinsonism. We targeted enrolment of 300 patients and 300 controls to assess feasibility and generate GP2-compatible genetic data. Recruitment continued beyond the target as additional sites became operational.

### Governance, ethics, and stakeholder engagement

The MoH, academic and clinical institutions, laboratories, patient groups, and community organisations contributed to regulatory procedures, biospecimen logistics, and community engagement. An institutional Memorandum of Understanding (MoU) between the MoH of the Kyrgyz Republic, KSMA, and the UzL supported implementation. The Kyrgyz Embassy in Germany facilitated institutional coordination and implementation. Ethical approval was obtained from the Bioethics Committee of KSMA (approval no. 3, Dec 27, 2023). Study protocols, informed consent documents, and data governance procedures were developed collaboratively with partners at the UzL and GP2. Local investigators retained responsibility for study conduct, participant engagement, and data stewardship.

### Research infrastructure and digital systems

Laboratory and biobanking infrastructure were established to support on-site sample processing and long-term storage. Standardised workflows for clinical assessment, biospecimen handling, data management, and governance were implemented and iteratively across participating sites (Supplementary Table S3). A secure local research database was developed to support pseudonymised management of clinical, environmental, and biospecimen data.

### Capacity development and training

Capacity development followed a tiered training model addressing clinical, laboratory, bioinformatics, and governance competencies (Supplementary Figure 1). Foundational training covered PD diagnosis, rating scales, biospecimen handling, research ethics, and introductory bioinformatics, supplemented by continuous online mentoring. Intermediate modules focused on genetics, data management, and governance, while advanced opportunities supported leadership development through specialist training and collaborative activities. In parallel, university curricula were strengthened by expanding instruction on extrapyramidal disorders and by integrating case-based learning.

### Participant recruitment and community engagement

Formative research, including focus-group discussions and stakeholder interviews, informed recruitment and engagement strategies. A Community Advisory Board comprising patients, caregivers, and clinicians advised on engagement materials and recruitment approaches. Participants were recruited through a university-affiliated movement disorders clinic in Bishkek established as part of the programme’s capacity-building activities, and through mobile outreach clinics across multiple regions. Participants enrolled in three groups: patients with PD, patients with PPS, and healthy controls. Eligible PD and PPS participants were aged ≥18 years, with diagnoses confirmed by a movement disorders specialist using international diagnostic criteria for PD, progressive supranuclear palsy, and multiple system atrophy^14–16^. Healthy controls were recruited through community outreach and recruitment activities based at participating hospitals and were screened to exclude parkinsonism and major neurological or psychiatric disorders. All participants provided written informed consent before enrolment.

### Clinical assessment and biospecimen processing

Participants underwent standardised clinical phenotyping, including clinical assessments and structured collection of environmental and lifestyle assessments using validated instruments (Supplementary Methods, Supplementary Tables 4, 5). The final versions of the study instruments and supporting documentation are available through the KyrPARK Zenodo repository. Participants consenting to longitudinal follow-up undergo repeat assessments at approximately 6-18 months, with long-term follow-up planned up to 10 years. Blood samples were processed according to standardised protocols and stored at −80 °C. DNA extraction, genotyping, and whole-genome sequencing (WGS), quality control, and data harmonisation were conducted and are ongoing according to established GP2 protocols and standards.^17,18^

### Statistical and data analysis

Baseline analyses were descriptive. Continuous variables are reported as mean with standard deviations (SD) or medians with interquartile ranges (IQR), as appropriate, and categorical variables as numbers (%). No formal hypothesis testing was performed. Feasibility and implementation metrics, including recruitment yield, data completeness, and biospecimen quality, were summarised descriptively. Analyses were based on available data for each variable, with no imputation of missing values; detailed patterns and reasons for missingness are provided in the Supplementary Methods.

For the initial genetic batch, variants in established PD-associated genes were extracted and analyzed using PLINK 1.9 and 2.0^19,20^, and annotated with a custom annotation pipeline based on ANNOVAR^21^. Additional findings from the genetic dataset have been reported previously.^8^ For descriptive reporting, variants comprised variants in established PD-associated genes meeting predefined relevance criteria, including pathogenic or likely pathogenic and variants with conflicting or uncertain interpretations of pathogenicity.

## Results

### Recruitment and participation

Between January 2024 and October 2025, we enrolled 1240 participants (549 with parkinsonism; 691 controls), reflecting the country’s rural–urban population structure and including individuals of Kyrgyz, Uzbek, Uyghur, Tatar, and other Central Asian ancestries, alongside smaller non–Central Asian minority groups. Baseline formative research (five focus-group discussions, n=30; 12 semi-structured interviews) identified limited awareness of PD and genetic research, alongside concerns about stigma, confidentiality, and data misuse. A 12-month public-engagement programme of clinician-led community meetings and regional radio broadcasts, alongside training in ethics, informed consent, and data governance, coincided with higher participation and a decline in refusal rates from 13·3% to 4·7%. Recruitment occurred through multiple clinical and community-based pathways (Table 1). Although sites in Bishkek accounted for the largest proportion of enrolment (56·9%), mobile and outreach-based recruitment strategies, including rural screening days, inpatient screening in non-neurological wards, occupational cohort screening, and a door-to-door prevalence pilot, contributed substantially (n=739). Recruitment via spouses and family networks contributed a relatively small number of controls (n=78). Participants were recruited from all seven administrative regions, with 58·5% residing in rural areas.

**Table 1.** Recruitment, coverage, and feasibility metrics (N=1240)

| Domain | Metric | Value |
| --- | --- | --- |
| **Recruitment infrastructure** | Active recruitment sites, n | 7 |
|  | Mobile outreach clinic deployments, n | 10 |
| **Recruitment sites** | University-affiliated movement disorders clinic, Bishkek, n (%) | 179 (14.4) |
|  | Clinical sites, Bishkek, n (%) | 526 (42.4) |
|  | Clinical sites, Osh, n (%) | 231 (18.6) |
|  | Focal points (rural Southern Kyrgyzstan), n (%) | 304 (24.5) |
| **Geographic coverage\*** | Regions represented, n (%) | 7/7 (100) |
|  | Regions with focal recruitment points, n (%) | 5/7 (71.4) |
| **Rural representation** | Participants from rural areas, n (%) | 726 (58.5) |
| **Recruitment modality\*\*** | University clinic and referral consultations, n (%) | 423 (34.1) |
|  | Mobile outreach clinics and other screening-based recruitment†, n (%) | 739 (59.6) |
|  | PD screening days in rural areas, n | 238 |
|  | Inpatient non-neurological screening, n | 346 |
|  | Other institution-based screenings, n | 70 |
|  | Door-to-door prevalence pilot, n | 85 |
|  | Spouses (healthy controls) and siblings (monogenic healthy controls), n (%) | 78 (6.3) |
| **Participation** | Eligible individuals with parkinsonism approached, n (%) | 602 (100) |
|  | Enrolled individuals with parkinsonism, n (%) | 549 (91.2) |
|  | Declined participation, n (%) | 53 (8.8) |

Percentages refer to enrolled participants unless otherwise specified.
\*Kyrgyzstan comprises 7 regions; “regions represented” includes participants from all regions, whereas “focal recruitment points” refer to regions with established recruitment sites.
\*\*Recruitment modality refers to the primary pathway through which participants were enrolled.
† Mobile outreach refers to discrete, organised screening activities and may include multiple encounters within the same region.
Eligible individuals with parkinsonism approached were those who met the inclusion criteria and were invited to participate. The proportion declining participation was calculated as the number of eligible individuals with parkinsonism who declined participation divided by the number of eligible individuals approached. Reasons for non-participation were not collected systematically across all sites. Where reported, reasons included limited familiarity with participation in research as a healthy control, particularly blood donation in the absence of perceived health need; concerns about genetic research or Parkinson’s disease–related stigma; and reluctance to provide blood samples.

### Participant characteristics

Baseline descriptive analyses included the first 300 patients with PD and 335 controls with complete baseline data (Table 2). Patients were older than controls (mean age 64·7 ± 9·6 vs 59·9 ± 11·2 years), whereas sex, ethnicity, and urban–rural distribution were broadly similar. 88·5% of participants reported Central Asian ancestry and 81·3% identified as Kyrgyz; parental consanguinity was uncommon. Educational attainment was modestly higher among controls. A PD family history was reported by 10·4% of patients. Self-reported pesticide exposure and prodromal features, including hyposmia and a positive RBD1Q screen, were more frequent among patients than controls. Additional baseline characteristics are summarised in Supplementary Table S6.

**Table 2.**
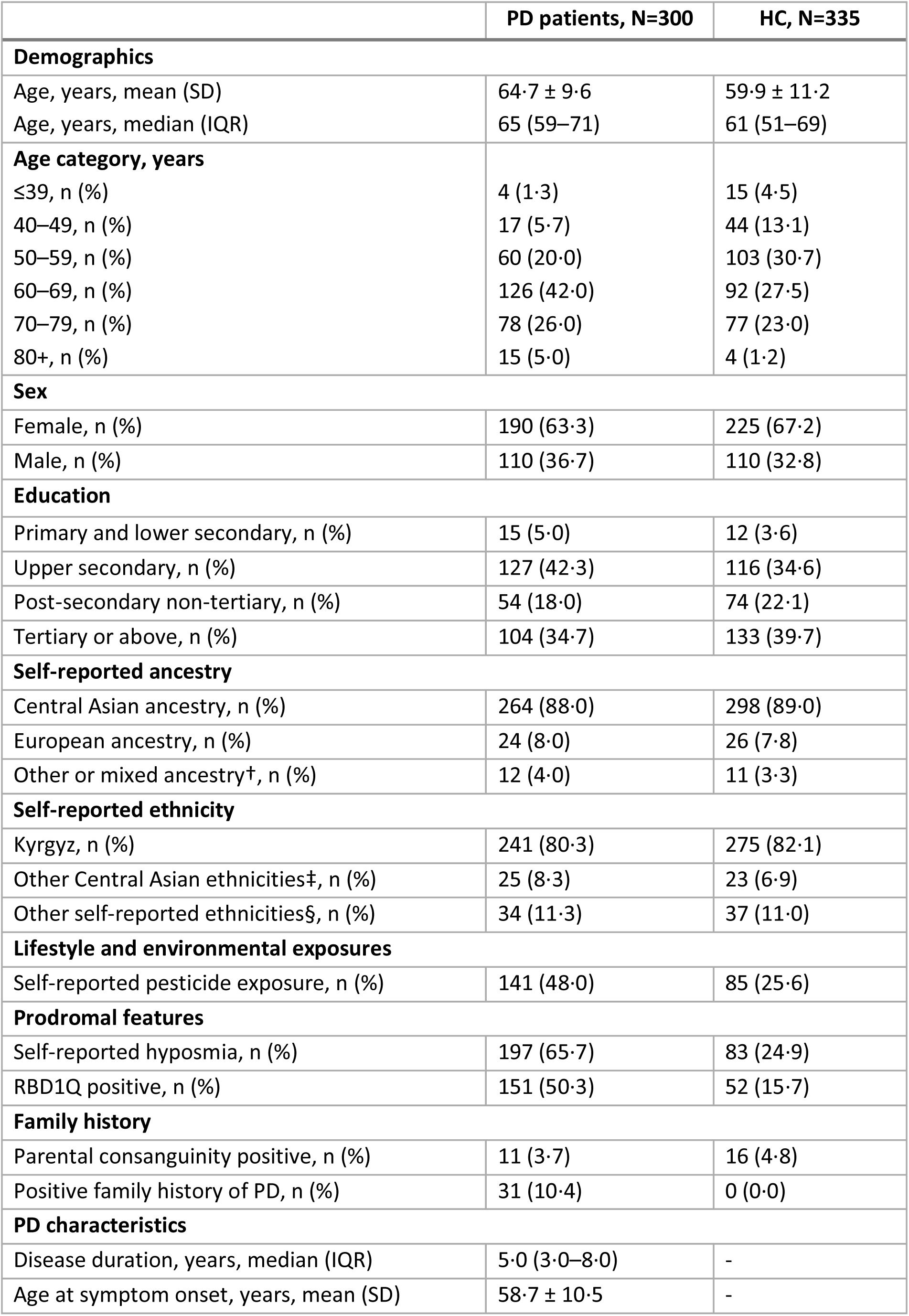
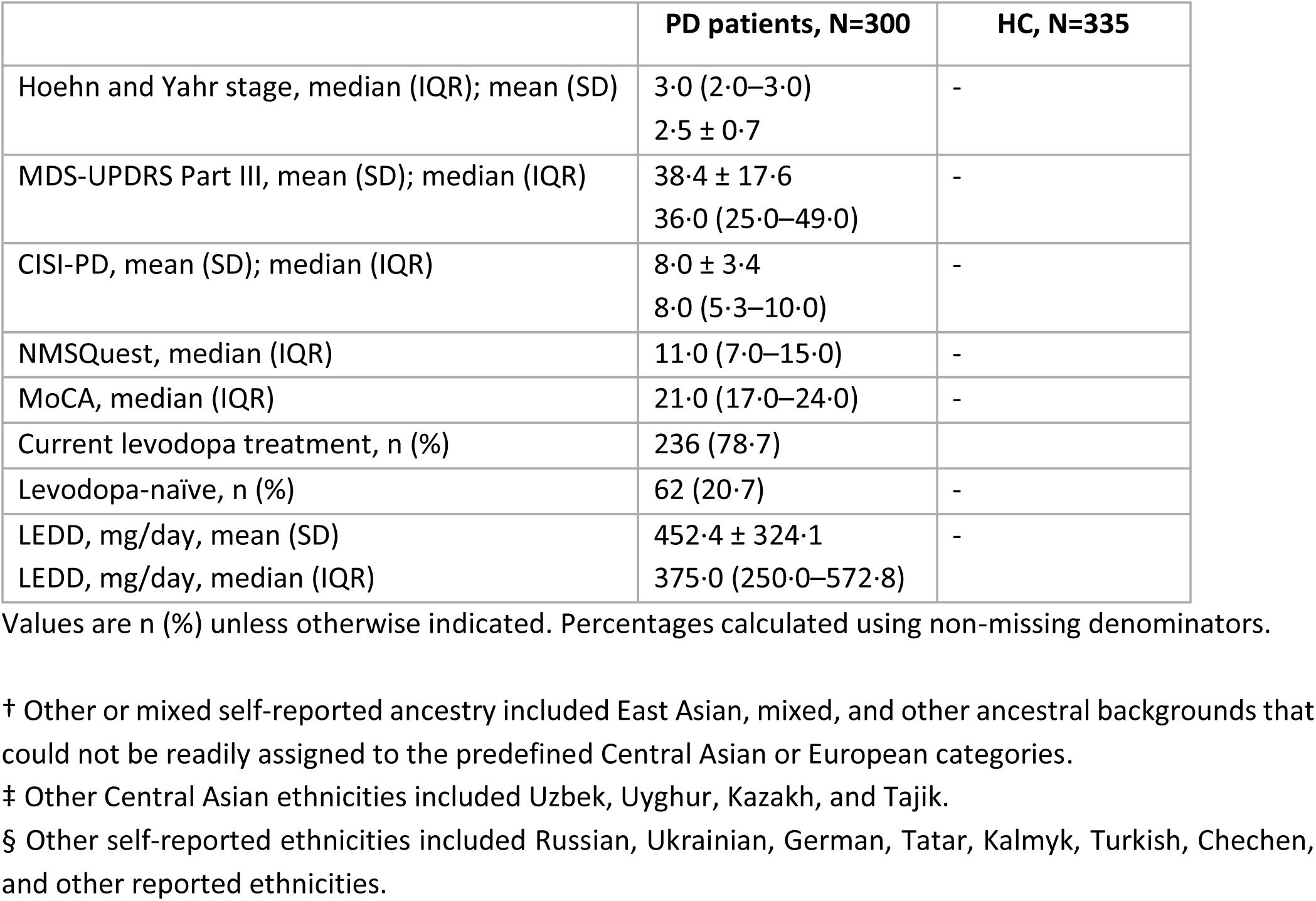
Baseline characteristics of the study cohort (N=635)

Among patients, median disease duration was 5·0 years (IQR 3·0–8·0), with a mean age at onset (AAO) of 58·7 years; 17·0% reported AAO before age 50 years. Median LEDD among treated patients was 375·0 mg/day (IQR 250·0–572·8; n=234). Median disease duration among the 62 levodopa-naïve participants was 3 years (IQR 2–6; range 0–16). NMSQest data were complete for 258 patients (86%), among whom the median non-motor symptom count was 11 (IQR 7–15); missingness was low overall, highest for the two sexual-function items (Supplementary Table S7).

### Biospecimen repository establishment and genomic data generation

Of the first 95 DNA samples processed, most met predefined quality thresholds on initial extraction, and a subset (n=7) required re-extraction. Genotyping followed GP2 quality-control standards, with samples shipped in batches to Novogene GmbH, Germany, for genotyping and WGS. Quality-control metrics for the initial batch are summarised in Supplementary Table S8; subsequent batches are ongoing. In the initial batch, self-reported sex was concordant with genetically inferred sex. Screening identified variants in a subset of patients (n=15/95; 15·8%) in established PD-associated genes (eg, *LRRK2* and *GBA1*), including variants classified as likely pathogenic and pathogenic, supporting the feasibility of downstream genetic analyses and inclusion of individuals from historically URP, including Kyrgyz and Uyghur participants (Supplementary Table S9).

### Workforce development and capacity strengthening

Between January 2024 and August 2025, 34 clinicians, four data scientists, two laboratory technicians, and additional research and support staff participated in a tiered training programme. Training was introduced in stages as the programme developed, beginning with research ethics and clinical phenotyping and later extending to laboratory methods, research methodology, data management, good scientific practice, and bioinformatics. Training combined in-person and online formats and included two workshops supported by the GP2 Training and Networking group,^22^ a hands-on laboratory training visit, and locally organised webinars, grand rounds, and needs-driven mentorship. A regional Central Asian movement disorders training camp convened clinicians and researchers and delivered hands-on instruction by local and international specialists, supported by the International Parkinson and Movement Disorder Society (MDS). MDS-UPDRS certification increased from three clinicians at baseline to 18 during the study period. Two clinicians received MDS–Asian and Oceanian Section visiting trainee grants, expanding the national movement disorders specialist pipeline in a setting that previously had a single trained specialist. Trainee-initiated subprojects were formalised as academic theses (n=3), grant applications were prepared (n=3), and abstracts were presented at international congresses (n=2). Participation expanded to include medical students (n=7), residents (n=6), and PhD students (n=5). Additional details are provided in Supplementary Table S10.

### Development of research infrastructure and collaborative networks

Implementation proceeded iteratively in two phases. Major implementation barriers, corresponding strategies, and subsequent adaptations are summarised in Figure 3. Clinical capacity expanded through the establishment of a university-based movement-disorders clinic and activation of six clinical sites across the country (Figure 1, Supplementary table S11), with undergraduate and postgraduate students participating in clinical and genetic research. A central biobank was established, alongside a secure local research database operating through a hybrid paper-based and electronic workflow. The partnership between the MoH, KSMA, and UzL was formalised through MoU. During the project, a national committee was established to review requests for international transfer of data and biospecimens, with a Data Sharing License required for each approved transfer.

**Figure 3.**
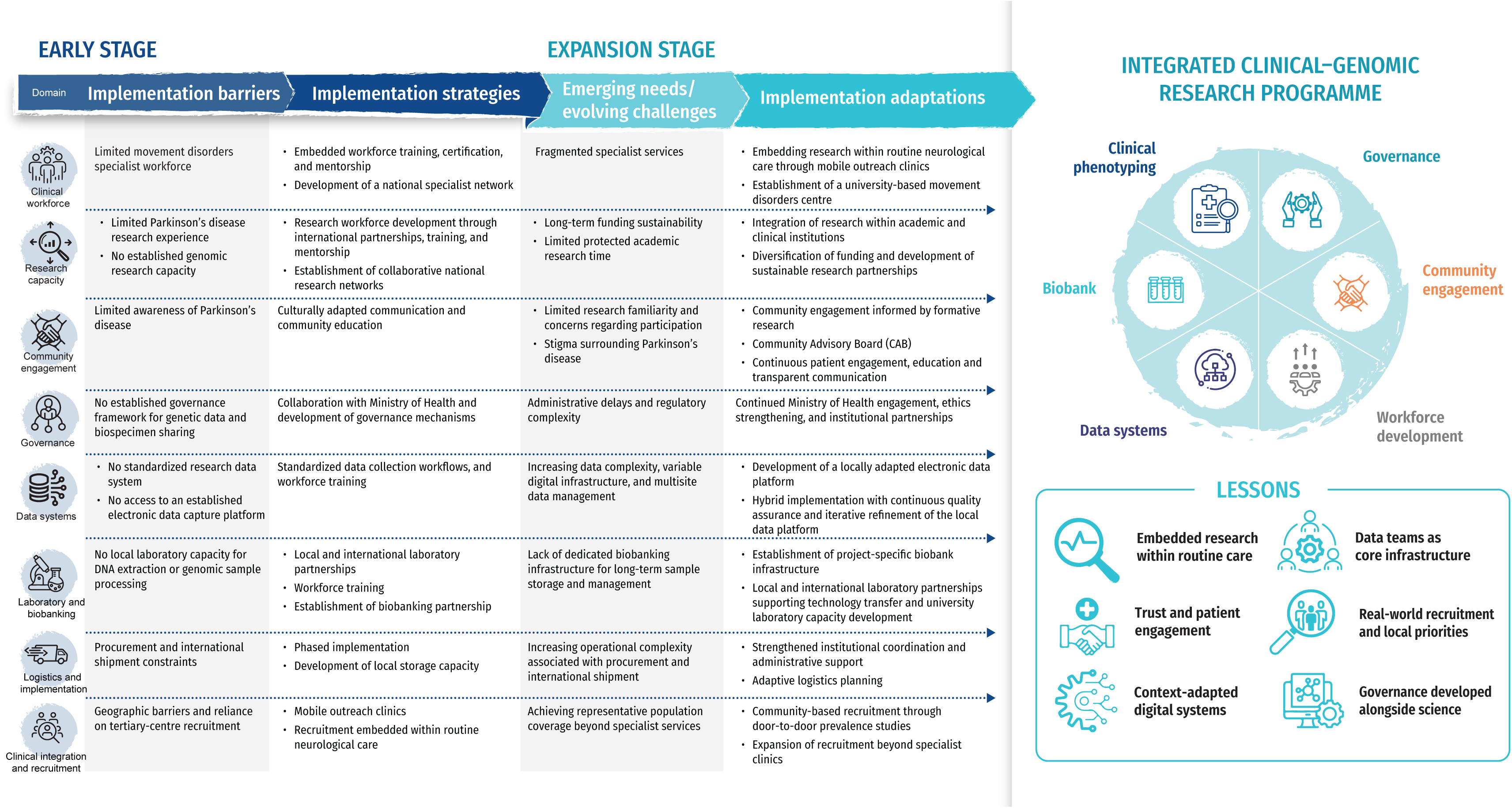
Context-adapted implementation strategies for establishing an integrated clinical–genomic Parkinson’s disease research programme in Kyrgyzstan. Early implementation barriers, implementation strategies, emerging challenges, and implementation adaptations during the establishment of a national Parkinson disease clinical–genomic research programme in Kyrgyzstan. The domains illustrate the core components of the integrated clinical– genomic research system developed through iterative implementation and capacity strengthening. The implementation principles represent transferable lessons derived from this experience.

## Discussion

Our findings suggest that embedding a structured capacity-building strategy within active research can facilitate the rapid establishment of an integrated clinical and neurogenetic infrastructure. Over 20 months, we developed a coordinated platform encompassing clinical phenotyping, biobanking, data governance, and community engagement in Kyrgyzstan, a country historically underrepresented in global neurogenomics (Figure 3). The genetics programme built on earlier locally initiated activities in training, patient engagement and education, and community-based support.^23^ Key implementation challenges included limited specialist capacity, the absence of established laboratory and data management systems, and the need to develop ethical and legal governance, while strengthening patient engagement and public awareness. Workforce development, digital systems, governance structures, and clinical and laboratory infrastructure evolved concurrently rather than sequentially, alongside expansion of clinical sites and increasing workforce certification. Progress was driven by aligning clinical, laboratory, and data systems with routine care rather than creating parallel research structures. In this setting, genomic research served as a catalyst for PD identification, standardised phenotyping, and registry-based surveillance.

Limited awareness of PD and persistent stigma, as reported in many LMIC settings, made formative research and community engagement integral throughout implementation. Close engagement with practicing clinicians, including those working in rural areas, together with patients and communities, helped ensure that implementation remained responsive to local priorities and realities and was embedded within routine clinical care.^23^ A recent survey of patients with PD in Kyrgyzstan identified trust in clinicians and researchers, clear communication, and continued patient engagement as important for research participation, while attitudes towards long-term biospecimen storage, recontact, and international data sharing were more cautious.^24^ Collaboration with trusted community structures, including religious and women-led forums, improved awareness of PD and genetic research, addressed concerns around stigma and confidentiality, and facilitated participation, consistent with experience from other LMIC genomics initiatives.^24–26^ Recruitment was more inclusive and efficient when delivered through mobile clinics and multidisciplinary outreach teams than through passive, tertiary-centre-based enrolment alone. Decentralised approaches reduced structural barriers, enabled real-time specialist consultation, integrated research with clinical care, and helped mitigate selection bias associated with referral-based recruitment. Our findings support decentralised recruitment as a feasible strategy in low-resource or geographically dispersed settings, particularly for PD, where under-ascertainment outside tertiary centres may influence observed disease patterns.^27,28^

Limited research experience and the absence of established data management systems in PD posed significant early challenges. Research training and data stewardship were embedded from the outset alongside development of a locally appropriate digital infrastructure to support data collection and quality assurance. A locally coordinated training initiative combined external training opportunities with regular webinars, mentorship, and peer learning within the research team. Iterative protocol refinement and targeted training helped standardise procedures across sites and accommodate varying levels of research experience. A phased transition from paper-based to hybrid electronic data capture provided an additional layer of quality assurance while clinical and research teams gained experience. In resource-limited settings, early investment in data management and analytical capacity may be as important as investment in clinical and laboratory infrastructure. Although some laboratory procedures can initially be supported through external collaborations, the capacity to generate, curate, harmonise, and interpret data locally is essential for maintaining data quality, scientific ownership, and long-term sustainability. Our experience suggests that digital infrastructure should be viewed as core research infrastructure rather than a secondary technical component.

As one of the first locally led human genetics research initiatives in the country, implementation required establishing procedures and governance mechanisms for international data and biospecimen sharing. Existing national experience from donor-supported programs for infectious diseases informed the development of local procedures for regulatory documentation and international data and biospecimen sharing. Early incorporation of broad consent and recontact permissions facilitated transition from cross-sectional enrolment to longitudinal follow-up without duplicative workflows.^3,18,29,30^ Close collaboration with the MoH and its affiliated agencies, as well as international partnerships, supported progress.

Many barriers became progressively easier to address once research activities began to strengthen clinical care and respond to locally identified priorities. Addressing unmet clinical needs and generating reliable data helped foster trust and sustained engagement among clinicians, patients, and institutions. Embedding governance alongside implementation reduced the need to redesign consent, data-sharing, and regulatory procedures. Together with broad consent and longitudinal follow-up, this infrastructure provides a platform for future biomarker, genetic, and gene–environment interaction studies.

### Strengths and limitations

A major strength of this study is the coordinated integration of clinical phenotyping, biobanking, digital data systems, governance, and community engagement within a nationally anchored framework, an uncommon achievement at an early stage in LMIC neurogenetics initiatives. Alignment with GP2 standards and MoH procedures supports methodological rigour, interoperability with international datasets, and the potential for long-term sustainability. The multiethnic composition and rural-urban coverage, including previously underrepresented Kyrgyz, Uyghur, and other Central Asian ancestries, address longstanding gaps in PD genomics. Genotype-based ancestry inference was used to account for population structure and enable investigation of ancestry-specific variants and cross-ancestry transferability. This work adds to the growing landscape of PD genetic research in Central Asia^30^ and to broader efforts to improve understanding of PD across URP.^30,32^

Several limitations warrant consideration. Individuals without contact with specialist or hospital services may remain underrepresented, potentially biasing estimates of disease burden and clinical characteristics. Controls were slightly younger than patients, reflecting pragmatic recruitment, and this difference should be considered when interpreting between-group comparisons. The cohort shows a higher proportion of tertiary-educated participants, and a female predominance likely reflects differential participation and health-seeking behaviour, although true sex-specific epidemiology cannot be excluded.^33^ Environmental exposure assessment currently relies primarily on self-reported data. Despite substantial gains in clinical, laboratory, and data-science capacity, workforce availability remains insufficient to meet national need, and long-term retention will depend on protected academic time, defined career pathways, and sustained institutional support. Dependence on external, project-based funding poses risks to long-term sustainability, and several governance and operational procedures have not yet been fully stress-tested at scale; formal evaluation of effectiveness and cost-effectiveness warrants dedicated assessment in subsequent phases.

### Scalability and transferability

The capacity-building model developed in Kyrgyzstan was intended to be scalable and transferable rather than exclusively context-specific. Core components likely to generalise across LMICs include early alignment with national health authorities, foundational governance structures, tiered workforce development, and decentralised recruitment integrated with service delivery. Scalability depends on adaptability rather than uniform replication, with community engagement, recruitment pathways, and training tailored to local social and health-system contexts. The staged implementation framework (Figure 2) and the synthesis of scalable programme components (Supplementary Table S1) distinguish core principles that are broadly transferable from implementation components that require adaptation to local contexts.

### Future Directions

Future work will consolidate this infrastructure into a sustainable research programme that integrates genomic, epidemiological, and clinical data. Recruitment will extend beyond specialist clinics into primary care and community based settings, with continued emphasis on rural and underserved populations. Generation of genotyping and WGS data, strengthened in country bioinformatics capacity, and family based designs will support ancestry informed and family analyses. More comprehensive environmental characterisation will be incorporated alongside population based and longitudinal studies, enabling integrated analyses of genetic and environmental determinants of PD in URP. Governance frameworks for data management, material transfer, and participant recontact will continue to evolve through implementation. As longitudinal data accrue, this infrastructure will support multicentre observational and interventional research through efficient participant recontact, harmonised phenotyping, and long term follow up.

## Conclusion

Our experience shows that neurogenetic research capacity can be established rapidly in resource limited settings through locally led, stepwise development, supported by sustained investment, institutional commitment, and international collaboration. By integrating clinical phenotyping, biobanking, data governance, workforce development, and community engagement within a nationally anchored framework, the Kyrgyzstan programme has established a platform for PD genetics and longitudinal research in populations historically underrepresented in genomic and PD research. This approach offers a pragmatic, adaptable model for advancing neurogenetic research and brain health capacity in other resource limited settings.

## Contributions

CS: Conceptualisation; Methodology; Investigation; Data curation; Formal analysis; Validation; Resources; Funding acquisition; Project administration; Supervision; Writing (original draft); Writing (review & editing).

EZ: Investigation; Data curation; Formal analysis; Project administration; Writing (review & editing).

AI: Investigation; Data curation; Project administration; Writing (review & editing).

MTP: Conceptualisation; Methodology; Writing (review & editing).

SK: Investigation; Data curation; Project administration; Writing (review).

CG: Formal analysis; Validation; Writing (review & editing).

SYL: Conceptualisation; Writing (review & editing). AK: Data curation; Writing (review).

AI [Iliazova], NB: Investigation; Data curation; Project administration; Writing (review).

GD: Conceptualisation; Resources; Project administration; Writing (review).

YK: Investigation; Project administration; Writing (review).

EP: Investigation; Data curation; Writing (review).

AM: Data curation; Project administration; Writing (review).

AS [Sorokin]: Formal analysis; Writing (review).

AA, SB [Bazarbaeva]: Investigation; Writing (review).

GP, BS: Investigation; Project administration; Writing (review).

AU, GZ, EZh: Investigation; Writing (review).

EM: Project administration; Writing (review & editing).

MB, AB [Baitelieva], AB [Bolotalieva], AT: Project administration; Writing (review).

JI: Investigation; Project administration; Writing (review).

NA: Investigation; Writing (review).

SB [Baltabaeva], AD, ZO: Investigation; Writing (review).

BN: Data curation; Project administration; Writing (review).

AM [Mukhanova]: Investigation; Writing (review).

IT: Resources; Project administration; Writing (review).

TK: Investigation; Writing (review & editing).

IS: Investigation; Writing (review). LML, JT: Writing (review & editing).

CB: Resources; Writing (review & editing).

AS [Singleton]: Resources; Writing (review & editing).

TFM: Resources; Project administration; Writing (review & editing).

AJN: Conceptualisation; Methodology; Resources; Supervision; Writing (review & editing).

IK: Conceptualisation; Resources; Project administration; Supervision; Writing (review & editing).

CK: Conceptualisation; Methodology; Resources; Funding acquisition; Supervision; Writing (original draft); Writing (review & editing).

CS and EZ directly accessed and verified the underlying study data. All authors reviewed and approved the final manuscript and accept responsibility for the decision to submit for publication. CS and CK had final responsibility for the decision to submit the manuscript.

## Data Availability

Data used in the preparation of this article were shared within the Global Parkinson’s Genetics Program (GP2; https://gp2.org). GP2 data can be accessed through AMP PD (https://amp-pd.org), subject to the relevant access requirements. The KyrPARK case-report form is publicly available in the KyrPARK Zenodo repository (https://doi.org/10.5281/zenodo.22011897). A data dictionary, study protocol, statistical analysis plan, and additional study materials are being prepared for deposition in the same repository.

## Code Availability

All code generated for this article, together with identifiers for the software programs and packages used, is available through the GP2code GitHub repository (https://github.com/GP2code/KyrPARK) and has been assigned a persistent identifier through Zenodo (https://doi.org/10.5281/zenodo.22261861).

## Acknowledgements

This project was supported by the Global Parkinson’s Genetics Program (GP2; https://gp2.org). GP2 is funded by the Aligning Science Across Parkinson’s (ASAP) (https://ror.org/03zj4c476) initiative and implemented by The Michael J. Fox Foundation for Parkinson’s Research (MJFF) (https://ror.org/03arq3225). For a complete list of GP2 members, see https://doi.org/10.5281/zenodo.7904831.

We acknowledge the MoH of the Kyrgyz Republic and the Embassy of the Kyrgyz Republic in Germany for institutional support and collaboration, the I.K. Akhunbaev Kyrgyz State Medical Academy (Bishkek, Kyrgyzstan) for hosting and facilitating programme activities, and the University of Luebeck for scientific collaboration and support.

We acknowledge GP2 Monogenic Network and GP2 Training and Networking Working Group for scientific and operational guidance and training opportunities during programme development and implementation.

We also acknowledge the Movement Disorders and Parkinson’s Disease Alliance Kyrgyzstan for its contribution to patient and community engagement and education. We thank all study participants, their families, and community members for their time, trust, and participation.

We also thank members of the Kyrgyzstan Parkinson’s disease research network for their contributions to programme implementation; a complete list is provided in Supplementary Table S11.

The authors used ChatGPT (OpenAI; GPT-5.6 Sol) and Perplexity Pro (Perplexity AI; GPT-5.6 Terra, Pro Search mode) to assist with language editing and improve clarity, readability, grammar, and sentence structure. These tools were not used to generate study data, conduct statistical analyses, interpret results, or draw scientific conclusions. All AI-assisted suggestions were reviewed and edited by the authors, who take full responsibility for the final manuscript.

