## Supplemental File for "A scalable model for Parkinson’s disease genetic research in underrepresented populations: lessons from Kyrgyzstan"

**Supplementary Appendix**

### Supplementary methods

#### Participant recruitment and eligibility

Recruitment for the analyses reported in this study occurred from 15 January 2024 to 30 September 2025. Detailed eligibility and exclusion criteria are reported in the main Methods. Controls were not individually matched to patients.

#### Clinical phenotyping

Standardised questionnaires captured demographic characteristics, lifestyle factors, environmental exposures, comorbidities, history of selected infectious diseases, residential history, family history of Parkinson's disease (PD) and other neurodegenerative disorders, and parental consanguinity. Demographic, lifestyle, environmental, residential, and family history variables were primarily based on participant self-report. Environmental exposures were assessed using the Mini Environmental Risk Questionnaire for Parkinson's disease (MERQ-PD; original and modified versions). Clinical assessments were performed by trained neurologists. Motor severity was assessed using the official Russian-language version of the Movement Disorder Society Unified Parkinson's Disease Rating Scale (MDS-UPDRS) Part III motor examination. Disease stage was assessed using the Hoehn and Yahr scale, and overall disease severity was assessed using the Clinical Impression of Severity Index for PD (CISI-PD). Non-motor symptoms were assessed using the Non-Motor Symptoms Questionnaire (NMSQuest), REM sleep behaviour disorder symptoms using the REM Sleep Behaviour Disorder Single-Question Screen (RBD1Q), and cognition using the Montreal Cognitive Assessment (MoCA). Hyposmia was assessed by self-report; the University of Pennsylvania Smell Identification Test (UPSIT) was introduced later in recruitment for a subset of participants. Additional assessments were introduced progressively as trained personnel and operational capacity became available and therefore were not administered to all participants at baseline. Assessments not administered because they had not yet been implemented were distinguished from participant non-response in the assessment of missing data. Medication use, dose, and dosing frequency were recorded during clinical assessment. Levodopa equivalent daily dose (LEDD) was calculated using established medication-specific conversion factors implemented through the Parkinson's Measurement LEDD Calculator.[1]

#### Demographic harmonisation

Educational attainment was recorded according to the Kyrgyz post-Soviet education system and subsequently harmonised to categories of the UNESCO International Standard Classification of Education (ISCED 2011)[2] to facilitate international comparisons (Supplementary Table S5). The original locally recorded educational categories were retained in the source dataset, and harmonised categories were derived for analysis. Ethnicity and ancestral background were self-reported using locally meaningful categories reflecting participants' identities and the regional context. Self-reported ethnicity and ancestry were treated as distinct from genetically inferred ancestry. For genetic analyses, ancestry was inferred from genotype-derived principal components and used for ancestry adjustment or ancestry-stratified analyses where appropriate.

**Data standardisation and quality assurance**

Standardised case report forms, variable definitions, and coding rules were developed before participant enrolment to promote consistency across study sites. Core study procedures were harmonised across sites, with subsequent refinement of selected forms and quality-control procedures as recruitment expanded. Clinical assessments followed standardised protocols, with investigator training and certification where required for individual instruments. Each participant was assigned a unique study identifier. This identifier was used to link clinical, questionnaire, and biospecimen data while maintaining pseudonymisation. Direct identifiers were managed separately from research datasets in accordance with study governance procedures.

Data quality was reviewed throughout recruitment using predefined quality-control procedures designed to identify missing values, implausible values, internal inconsistencies, duplicate or incompatible coding, and incomplete records. Queries generated during quality control were communicated to recruiting sites and, where appropriate, prompted source-data verification and correction of study records. Recurrent problems were addressed through targeted feedback, clarification of coding rules, or retraining of study personnel. As recruitment expanded to additional sites, quality-control procedures were iteratively refined to address recurring data-quality issues while maintaining consistent variable definitions and coding conventions across the study. Corrections were made against available source documentation rather than inferred from other variables.

Before transfer, variables were harmonised to GP2 data standards, including variable naming, coding, and formatting. Revised datasets underwent further quality-control checks before submission to the GP2 platform, and quality-control reports were shared with the GP2 coordinating team as part of the harmonisation process.

**Missing data**

Missingness was assessed for each variable and summarised by variable domain (Supplementary Table S11). Missing values included participant non-response, including responses recorded as “prefer not to answer” or “I don’t know”, and assessments or questionnaire items that were not administered. These categories were distinguished where source data permitted. Some variables had higher levels of missingness because the corresponding assessments were introduced after recruitment had begun or were implemented in phases. Missingness for these variables therefore reflects study implementation as well as participant-level non-response and should not be interpreted solely as loss of collected data. Analyses were based on available data for each variable, using variable-specific denominators. No imputation of missing values was performed unless otherwise specified. For categorical variables, percentages were calculated using the relevant non-missing, group-specific denominator unless otherwise stated.

**Measures to support consistency and reduce bias**

Several procedures were used to promote consistent ascertainment across recruitment settings. Diagnoses of PD and PPS were confirmed by a movement disorders specialist using internationally recognised diagnostic criteria; healthy controls were screened for parkinsonism and major neurological or psychiatric disorders; clinical assessments were conducted using standardised instruments and harmonised study procedures; and participating investigators received study-specific training and certification where applicable (Methods section).

Recruitment through both clinic-based and outreach pathways was used to extend recruitment beyond a single tertiary-care setting. Nevertheless, recruitment was not population-based, and differences in referral, health-care access, willingness to participate, or outreach coverage could have influenced the characteristics of enrolled participants. These limitations are considered in the Discussion.

**Supplementary Table S1. Characteristics of publications included in the evidence synthesis**

| First author, year | Journal | Scope* | Setting ** | Study type | Primary contribution | Extent of implementation description† | Implementation domain(s) described | Main relevance to this study |
|---|---|---|---|---|---|---|---|---|
| Nalls MA et al., 2019 | Lancet Neurol | Global (predominantly European ancestry) | Mixed | GWAS meta-analysis | Identified ~90 independent PD risk loci; the largest pre-GP2 PD GWAS meta-analysis and the historical diversity-gap benchmark. | None | — | Demonstrates the scientific value of ancestry-diverse PD genetics and provides a discovery comparator. |
| Kim JJ et al., 2024 | Nat Genet | Global | Mixed | GWAS meta-analysis | First large-scale multi-ancestry PD GWAS; identifies ancestry-shared and ancestry-specific loci via the GP2. | None | — | Demonstrates the scientific value of ancestry-diverse PD genetics and provides a discovery comparator. |
| Rizig M et al., 2023 | Lancet Neurol | Africa and African ancestry | Mixed | Original research (GWAS) | First large African-ancestry PD GWAS; identifies novel loci including a common GBA1 risk variant. | Limited | Regional collaboration; cohort recruitment | Demonstrates the scientific value of ancestry-diverse PD genetics and provides a discovery comparator. |
| Jonson C et al., 2024 | Alzheimers Dement | Global | Mixed | Systematic review | Quantifies persistent lack of ancestral diversity across neurodegenerative-disease GWAS in the GWAS Catalog and literature. | None | — | Provides structured evidence on underrepresentation or a regional PD genetics landscape. |
| Lim SY et al., 2024 | Lancet Neurol | Global | Mixed | Narrative review/ perspective | Synthesises global PD genetic discoveries and their path to clinical translation, highlighting international/LMIC capacity gaps. | Moderate | Workforce and research capacity; partnerships; governance; clinical translation | Links global PD genetics to translational and capacity needs relevant to the implementation model. |
| Santos-Lobato BL et al., 2021 | Arq Neuropsiquiatr | Brazil | LMIC | Systematic review | Systematically reviews monogenic PD genetic forms reported in Brazil. | None | — | Provides structured evidence on underrepresentation or a regional PD genetics landscape |

| | | | | | | | | |
|---|---|---|---|---|---|---|---|---|
| Schumacher-Schuh AF et al., 2022 | Mov Disord | Global | Mixed | Systematic review | Maps the current landscape of underrepresented populations in PD genetics research and proposes future directions. | Limited | Strategic priorities; partnerships; workforce and research capacity; governance | Defines the representation gap and regional evidence base underpinning the study rationale. |
| Mohamed W et al., 2023 | Am J Neurodegener Dis | Africa and Arabia | Mixed | Narrative review | Reviews the monogenic PD genetic-diversity landscape and research gaps in the AfrAbia region. | Limited | Strategic priorities; regional research capacity | Defines the representation gap and regional evidence base underpinning the study rationale. |
| Khani M et al., 2024 | Ann Neurol | Global | Mixed | Perspective | Proposes a strategic framework for globalizing PD genetics research and expanding representation. | Limited to Moderate | Partnerships; governance and strategic planning; sustainability | Identifies strategic priorities for global PD genetics but provides limited operational implementation evidence. |
| Marconi GA et al., 2025 | Curr Opin Neurol | Global | Mixed | Narrative review | Summarises recent advances in PD genetics research specifically in underrepresented populations. | Limited | Strategic priorities for inclusion of underrepresented populations | Defines the representation gap and regional evidence base underpinning the study rationale. |
| Blanckenberg J et al., 2013 | J Neurol Sci | Sub-Saharan Africa | LMIC | Narrative review | Early synthesis of PD prevalence and genetics knowledge (and gaps) in sub-Saharan Africa. | Limited | Regional research gaps; future research priorities | Defines the representation gap and regional evidence base underpinning the study rationale. |
| Mohamed W et al., 2024 | J Integr Neurosci | Africa and Arabia | Mixed | Consortium programme review | Describes the AA-PD-GC consortium's composition, activities and future perspectives for PD genetics in the AfrAbia population. | Moderate-to-extensive | Consortium and network development; partnerships; governance and strategic planning | Closest regional programme comparator; informs network, partnership, and capacity-building discussion. |
| Arias-Carrión O et al., 2026 | Front Aging Neurosci | Mexico | LMIC | Systematic review | Systematically reviews PD genetic architecture literature in Mexico. | None | — | Provides structured evidence on underrepresentation or a regional PD genetics landscape. |

| | | | | | | | | |
|---|---|---|---|---|---|---|---|---|
| Flores-Ocampo V. et al., 2026 | J Parkinsons Dis | Global | Mixed | Consortium programme review | Comprehensive overview of GP2's design, scale, and capacity-building activities since 2020 | Extensive | Governance; funding/partnerships; workforce and capacity building; data systems; biobank/sample collection | Describes funding structure (ASAP/MJFF), data-sharing model, training/fellowship programs, and cohort integration. |
| Blauwendraat C. et al., 2025 | Am J Hum Genet | Global | Mixed | Perspective/roadmap | Roadmap detailing how GP2 operationalises equity, capacity building, and a sustainable research collective. | Moderate-to-extensive | Governance; sustainability; capacity building; partnerships | Programmatic roadmap with named operational principles (equity, capacity, sustainability) |
| Singleton A. et al., 2020 | J Parkinsons Dis | Global | Mixed | Consortium programme review | Retrospective on IPDGC's 2009 founding, growth to 100+ scientists, and evolving research focus over its first decade. | Moderate | Consortium/ network development; partnerships; governance | Describes founding, informal trust-based governance model, and growth, but not detailed operational metrics. |
| Vollstedt E.-J. et al., 2023 | PLOS One | Global | Mixed | Empirical implementation study | Describes building and operating an online resource (MJFF Global Genetic PD Project) to collect and share monogenic PD case data from underrepresented populations worldwide. | Extensive | Data systems (online platform); community/ global collaboration; underrepresented-population inclusion | Reports the platform's build process, data-collection workflow, and inclusion strategy as primary content. |
| Bandres-Ciga S. et al., 2024 | Mov Disord | Global, multi-ancestry | Mixed | Platform development | Describes design and validation of the NeuroBooster Array, a genotyping platform built for multi-ancestry neurological-disease genetics. | Extensive | Data systems / genotyping platform | Reports the actual array design, variant content, and validation across populations. |

*Region refers to the primary geographic scope of the study population or programme described. ** Setting refers to the research and implementation context in which the study was conducted or the programme operated (low- and middle-income countries (LMIC), high-income countries (HIC), or multinational mixed settings), rather than solely the ancestry or country of origin of participants. Study type reflects the primary objective and design of the publication. † Implementation focus was classified according to the extent to which the publication described the establishment, governance, operationalisation, or evaluation of research infrastructure or programmes: None, no description of implementation or research infrastructure; Limited, implementation mentioned as background, rationale, or future direction; Moderate, selected implementation components (e.g., partnerships, governance, workforce, capacity building, or consortium organisation) described but implementation was not the primary focus; Extensive, implementation, programme development, infrastructure, governance, or operational processes constituted the primary focus of the publication. Implementation domains were assigned inductively based on the principal implementation components explicitly described in each publication.

**Supplementary Table S2. Scalable and context-adaptable components of the capacity-building framework**

| Domain | Scalable component | Rationale for scalability | Context-specific adaptations |
|---|---|---|---|
| Governance & alignment | Early alignment with Ministry of Health and key universities | Working through national health authorities and academic hubs is a generalizable way to anchor neurogenetic research and ensure legitimacy. | Which ministry/agency leads, specific universities or hospitals involved, and how formal the agreements are. |
| Regulatory, ethics, and data/material governance | Early engagement with regulators and co-developed ethics, consent, and data/material governance with local and international partners | The "minimum viable" governance structures and co-development of protocols are process templates that can be reused in other LMICs. | National regulatory and ethics frameworks, consent language, data-protection and export laws, and positions on data and biospecimen transfer and return. |
| Capacity- building framework | Seven checkpoint, question-based capacity-building framework (institutional alignment, governance, infrastructure, workforce, engagement, partnerships, sustainability) | This is explicitly presented as a generic implementation scaffold rather than a context-specific protocol and is supported by illustrative indicators and adaptable examples. | Which checkpoints are prioritized first, speed of progression, types of partners and funders. |
| Clinical model | Multisite case–control cohort with an embedded longitudinal component | A tiered design (case–control plus longitudinal follow-up) is widely applicable for PD genetics and other neurodegenerative conditions. | Disease focus, recruitment targets, follow-up schedule, and specific assessments used. |
| Service-linked recruitment | Use of university-based movement-disorder clinics plus mobile outreach clinics | Coupling research with service delivery and mobile outreach is a replicable strategy to include rural and underserved populations. | Whether outreach is delivered through mobile vans, satellite clinics, or telemedicine, depending on geography, transport, and local safety considerations. |
| Community engagement | Community Advisory Board, formative qualitative work, multilingual materials, media and community-leader engagement | An engagement package combining CAB input, formative research, and locally tailored materials can be applied in many settings. | Composition of CAB, languages, media channels, cultural tailoring of messages. |
| Digital infrastructure | Locally developed electronic data capture (EDC) platform with role-based access and parallel paper workflows | The principle of a secure, locally hosted EDC that complies with national data laws and mirrors paper workflows is generalizable. | Specific software, hosting solution (local server vs cloud), and integration with national systems. |
| Biobanking workflows | Standardized SOPs for sample collection, processing, storage, and shipment | SOP-based biobanking that is aligned with GP2 standards can be transplanted to other centers with appropriate training. | Available equipment, storage capacity, and shipping options. |
| Training model | Tiered, competency-based training (foundational, intermediate, advanced) | A tiered training ladder that links short courses, mentorship, and curriculum reform is broadly adaptable. | Content focus (e.g., which diseases, which omics), duration, and mode (in-person vs online). |

| | | | |
|---|---|---|---|
| | with ongoing mentorship and curriculum integration | | |
| Partnership model | Integration into GP2 and support from ASAP/MJFF; shared protocols and data standards | Joining a global genetics consortium and using shared standards is a scalable partnership blueprint for other regions. | Which consortium(s) are joined and what level of data/sample sharing is feasible under local law. |
| Monitoring and adaptive learning | Embedded monitoring and feedback loops across implementation domains | Continuous monitoring supports adaptive implementation and early identification of bottlenecks. | Availability of local monitoring systems, reporting requirements, and human resources. |
| Scalability philosophy | Focus on functional requirements rather than fixed institutional forms | The explicit emphasis on adaptability and defining functions (what must exist) rather than structures (what it must look like) is inherently transferable. | Concrete institutional arrangements will differ, but the "functional requirements" framing remains constant. |

* Components are interdependent and intended as a generalisable implementation scaffold requiring local adaptation.

**Supplementary Table S3. Overview of standardised research workflows**

| Domain | Workflow component | Tools | Purpose |
|---|---|---|---|
| Governance | Consent, access review | Consent workflow<br>Export document preparation workflow | Ethical compliance |
| Clinical | Participant assessment & diagnosis | Clinical data collection protocol based on the study branch (PD, PPS, Monogenic, HC)<br>MDS-UPDRS, NMSQuest, TUG test, MoCA<br>MERQ-PD, Modified MERQ-PD<br>Video/audio recording protocols | Standardised phenotyping |
| Biospecimens | Blood collection, processing, storage | Blood collection and cold-chain in-country transportation protocol<br>Blood sample preparation and storage protocol<br>DNA extraction from blood (protocol)<br>DNA storage protocol<br>Sample local QC protocol<br>Transportation protocol | Sample integrity |
| Data | De-identification & EDC entry | Data de-identification workflow<br>Data entry workflow<br>Data entry QC workflow<br>Standardisation and QC | Data security & completeness |
| | Data harmonisation and transfer | Ethical and legal compliance<br>Cross-site compatibility<br>Data preparation and harmonisation workflow<br>Data QC workflow | Data harmonization & integration |

*Workflows were standardised across sites in alignment with international research standards and adapted as needed to local regulatory and operational contexts.

**Supplementary Table S4. Standardised clinical assessments and data collected within the Kyrgyzstan Parkinson's disease research programme**

| Domain | Assessment/instrument | Participants | Baseline | Follow-up |
|---|---|---|---|---|
| Demographics | Demographic questionnaire | All | ✓ | Updated as applicable |
| Education | Harmonised ISCED categories | All | ✓ | NA |
| Ethnicity | Self-reported ethnicity | All | ✓ | NA |
| Ancestry | Self-reported ancestral background | All | ✓ | NA |
| Genetic ancestry | Genotype-derived principal components | Genotyped subset | ✓ | NA |
| Family history and consanguinity | Structured study questionnaire | All | ✓ | Updated as applicable |
| Disease history | Standardised study questionnaire (age at onset, disease duration, diagnostic history, symptoms, medication history) | PD/PPS | ✓ | Updated as applicable |
| Clinical diagnosis | MDS Clinical Diagnostic Criteria for PD / MDS Diagnostic Criteria for PSP and MSA | PD/PPS | ✓ | Updated as applicable |
| Motor severity | MDS-UPDRS Part III | PD/PPS | ✓ | ✓ |
| Disease stage | Hoehn and Yahr stage | PD/PPS | ✓ | ✓ |
| Functional mobility | Timed Up and Go (TUG) test* | PD/PPS | ✓ | ✓ |
| Disease severity | Clinical Impression of Severity Index for PD (CISI-PD) | PD | ✓ | ✓ |
| Cognition | Montreal Cognitive Assessment (MoCA) | PD/PPS | ✓ | ✓ |
| Non-motor symptoms | Non-Motor Symptoms Questionnaire (NMSQuest) | PD/PPS | ✓ | ✓ |
| REM sleep behaviour disorder | RBD Single-Question Screen (RBD1Q) | All | ✓ | ✓ |
| Hyposmia | Self-report | All | ✓ | Updated as applicable |
| Olfaction | University of Pennsylvania Smell Identification Test (UPSIT)* | Subset | ✓ | NA |
| Environmental exposures | MERQ-PD (original and modified versions) | All | ✓ | ✓ |
| Residential history | Structured study questionnaire | All | ✓ | Updated as applicable |
| Lifestyle factors | Structured study questionnaire | All | ✓ | ✓ |
| Comorbidities | Structured study questionnaire | All | ✓ | Updated as applicable |
| Medication | Antiparkinsonian medication inventory | PD/PPS | ✓ | ✓ |
| Treatment burden | Levodopa equivalent daily dose (LEDD) | PD/PPS | ✓ | ✓ |
| Biospecimens | Whole blood and DNA | All | ✓ | Selected participants |

*Introduced during later phases of recruitment and therefore not available for all participants at baseline. PD=Parkinson's Disease; PPS=Parkinsonian syndromes; HC=healthy controls. NA=not applicable. "All" denotes participants across the study groups for whom the relevant assessment was applicable.

**Supplementary Table S5. Harmonisation of educational attainment from the Kyrgyz or Soviet education system to UNESCO International Standard Classification of Education (ISCED 2011) categories**

| Local category | Harmonised category | ISCED level |
|---|---|---|
| No formal education, Primary education | Primary education or less | 0–1 |
| Incomplete secondary education | Lower secondary education | 2 |
| Complete secondary education | Upper secondary education | 3 |
| Vocational/professional and secondary specialized education | Post-secondary non-tertiary education | 4 |
| Higher education (Specialist degree, Master's degree) | Tertiary education | 6–7 |
| Postgraduate degree (PhD, Candidate of Sciences, Doctor of Sciences) | Doctoral education | 8 |

Educational attainment was harmonised according to the UNESCO International Standard Classification of Education (ISCED 2011) to facilitate international comparability and pooled analyses across cohorts.

**Supplementary Table S6. Comorbidities and lifestyle factors (N=635)**

| | PD patients, N=300 | HC, N=335 |
|---|---|---|
| **Comorbidities** | | |
| Hypertension, n (%) | 146 (49·0) | 156 (46·8) |
| Type 2 diabetes, n (%) | 26 (8·7) | 50 (14·9) |
| Cardiovascular disease, n (%) | 39 (13·1) | 56 (16·8) |
| Depression, n (%) | 214 (71·3) | 64 (19·3) |
| **Lifestyle factors and history** | | |
| Ever smoked, n (%) | 59 (19·7) | 83 (24·8) |
| Alcohol use, n (%) | 54 (18·1) | 77 (23·0) |
| Head trauma, n (%) | 84 (28·2) | 96 (28·7) |
| Nasway use (ever/current), n (%)† | 9 (4·2) | 11 (3·3) |

Percentages were calculated using variable-specific non-missing denominators. Missingness is summarised in Supplementary Table S11. † Nasway is a locally used form of smokeless tobacco, typically placed in the oral cavity. The nasway item was introduced after study initiation.

**Supplementary Table S7. Non-motor symptoms reported on the NMSQuest among patients with Parkinson's disease (N=300)**

| NMSQuest item | Responded, n | Symptom present, n (%) | Missing, n (%) |
|---|---|---|---|
| Nocturia | 299 | 195 (65·2%) | 1 (0·3%) |
| Depression | 297 | 193 (65·0%) | 3 (1·0%) |
| Constipation | 300 | 180 (60·0%) | 0 |
| Restless legs syndrome | 297 | 178 (59·9%) | 3 (1·0%) |
| Anxiety | 300 | 177 (59·0%) | 0 |
| Subjective cognitive complaint | 298 | 170 (57·0%) | 2 (0·7%) |
| Insomnia | 300 | 163 (54·3%) | 0 |
| Subjective orthostatic symptoms | 300 | 157 (52·3%) | 0 |
| Pain | 300 | 147 (49·0%) | 0 |
| Urinary urgency | 299 | 143 (47·8%) | 1 (0·3%) |
| Hyposmia | 299 | 137 (45·8%) | 1 (0·3%) |
| Incomplete bowel emptying | 299 | 123 (41·1%) | 1 (0·3%) |
| Sweating | 296 | 116 (39·2%) | 4 (1·3%) |
| Attention difficulty | 297 | 115 (38·7%) | 3 (1·0%) |
| REM sleep behaviour disorder symptoms | 300 | 116 (38·7%) | 0 |
| Vivid dreams | 298 | 112 (37·6%) | 2 (0·7%) |
| Leg swelling | 296 | 98 (33·1%) | 4 (1·3%) |
| Reduced sexual interest | 277 | 90 (32·5%) | 23 (7·7%) |
| Sexual difficulty | 267 | 85 (31·8%) | 33 (11·0%) |
| Excessive daytime sleepiness | 300 | 84 (28·0%) | 0 |
| Dysphagia | 299 | 79 (26·4%) | 1 (0·3%) |
| Unintentional weight loss | 300 | 76 (25·3%) | 0 |
| Falls | 300 | 74 (24·7%) | 0 |
| Sialorrhoea | 299 | 69 (23·1%) | 1 (0·3%) |
| Nausea | 299 | 50 (16·7%) | 1 (0·3%) |
| Visual hallucinations | 299 | 47 (15·7%) | 1 (0·3%) |
| Diplopia | 298 | 41 (13·8%) | 2 (0·7%) |
| Delusions | 296 | 33 (11·1%) | 4 (1·3%) |
| Faecal incontinence | 298 | 23 (7·7%) | 2 (0·7%) |

Percentages for individual symptoms are calculated among participants who responded to the respective item. Missing responses were retained as missing and were not classified as symptom absence. NMSQuest total symptom count was calculated among participants with complete responses to all 30 items.

**Supplementary Table S8. Quality-control metrics and sample flow for the initial batch of samples for genetic analysis (N=95)**

| QC domain | Metric | Value |
|---|---|---|
| Samples flow | Samples extracted and shipped (batch 1) | 95 |
| | Samples excluded before genotyping (with reason) | 4 |
| | Samples entering the genetic analysis QC pipeline | 91 |
| | Samples excluded after QC (total) | 3 |
| | Samples passing QC and released for analysis | 88 |
| Genotyping performance | Mean call rate (%) | 96·2% |
| Sample concordance | Sex concordance (recorded vs genetically inferred) | 88/88 (100%) |
| Population structure | Samples with genetically inferred ancestry assigned | 88/88 (100%) |
| | Concordance between self-reported ancestry and genetically inferred ancestry | 86/88 (97·7%) |
| | Discordant ancestry assignments identified* | 2/88 (2·3%) |

* Genetically inferred ancestry was based on PCA. Most participants self-identifying as Kyrgyz, Uzbek, and Uyghur clustered within Central Asian ancestry, while the participant self-identifying as Korean clustered within East Asian ancestry, and most participants self-identifying as Russian, Ukrainian, and German clustered within European ancestry. Two participants self-identifying as Tatar clustered within European genetic ancestry; these discordant ancestry assignments were reviewed and showed no evidence of sample mix-up or quality-control failure.

**Supplementary Table S9. Descriptive summary of PD-related variants identified among patients with Parkinson's disease in the initial genetic analysis batch**

| Gene | Genomic variant (GRCh38) | Amino acid change | rsID | Genetically inferred ancestry | PD carriers, n (zygosity) | Analytical source |
|---|---|---|---|---|---|---|
| *GBA1* | chr1:155236376:C:T | p.Glu365Lys | rs2230288 | CAS (3); EUR (1) | 4 (HET) ‡ | PLINK/ANNOVAR analysis |
| *GBA1* | chr1:155236246:G:A | p.Thr408Met | rs75548401 | EUR | 2 (HET) | PLINK/ANNOVAR analysis |
| *GBA1* | chr1:155239936:C:T | p.Arg86Gln | rs144173415 | CAS | 2 (HET) | PLINK/ANNOVAR analysis |
| *GBA1* | chr1:155238210:C:T | p.Ala229Thr | - | CAS | 1 (HET) | WGS |
| *GBA1* | chr1:155237524:T:G | p.Glu272Asp | - | CAS | 1 (HET) | WGS |
| *GBA1* | chr1:155237412:T:C | p.Ser310Gly | rs1057942 | CAS | 1 (HOM) | PLINK/ANNOVAR analysis |
| *GBA1* | chr1:155235843:T:C | p.Asn409Ser | rs76763715 | EUR | 1 (HET) | PLINK/ANNOVAR analysis |
| *GBA1* | chr1:155235252:A:G | p.Leu483Pro | rs421016 | CAS | 1 (HET) ‡ | WGS (Gauchian) |
| *GBA1* | - | c.1263del+RecTL | - | CAS | 1 (HET) | WGS (Gauchian) |
| *LRRK2* | chr12:40252984:C:T | p.Ala419Val | rs34594498 | CAS | 1 (HET) | PLINK/ANNOVAR analysis |
| *LRRK2* | chr12:40320043:G:C | p.Arg1628Pro | rs33949390 | CAS | 1 (HET) | PLINK/ANNOVAR analysis |

This table provides a descriptive summary of variants identified in the initial genetic analysis batch and should not be interpreted as an estimate of variant frequency or genetic association. Comprehensive genetic analyses will be reported separately. CAS = Central Asian genetic ancestry (PD patients self-identifying as Kyrgyz or Uyghur, with genetically inferred Central Asian ancestry); EUR = European genetic ancestry (PD patients self-identifying as Russian or Ukrainian, with genetically inferred European ancestry). HET = heterozygous; HOM = homozygous. WGS = whole-genome sequencing. ‡One participant carried both *GBA1* p.Glu365Lys and p.Leu483Pro.

**Supplementary Table S10. Overview of training activities, participation, and early outputs**

| Training domain | Activity / modality | Participants (n) | Roles represented | Outputs / status |
|---|---|---|---|---|
| Clinical training | PD diagnosis, phenotyping, and hands-on movement disorders training (1st GP2 workshop, MDS-supported courses and camps, local webinars, and hands-on training) | 34 | Neurologists, residents | Completed |
| | Grand rounds, online seminars (local) | 35 | Neurologists, residents | Ongoing |
| | MDS-UPDRS certification (online; MDS web-based training) | 18 | Neurologists | Completed |
| Laboratory training | Biospecimen collection & processing, and SOP implementation (hands-on training by UzL, GP2 workshops, and local hands-on training) | 5 | Laboratory staff, clinicians | Completed/ Ongoing |
| Data & bioinformatics | Bioinformatics GP2 workshops and GP2 Learning Platform | 16-30 (activity-dependent) † | Clinicians, Data scientists | Completed/ Ongoing |
| Research methods, ethics and good scientific practice | Research methods modules (1st GP2 workshop) | 30 | Clinicians, PhD students | Completed |
| | Research ethics and responsible conduct of research (GP2 workshops, local hands-on training) | 16-34 (activity-dependent) † | Clinicians, Data scientists, Core research team/personnel | Completed/ Ongoing |
| | Applied statistics and clinical research data analysis (local) | 7 | Core research team/personnel; Clinician-scientists | Ongoing |
| | Good Scientific Practice seminars and workshops (local) | 7 | Core research team/personnel; Clinician-scientists | Ongoing |
| Mentorship & leadership | Project-based mentorship | 3 | Trainees | Ongoing |
| Early outputs | Trainee-led research subprojects | 3 | Clinician-scientists | Formalised as theses (PhD) |
| | Abstracts submitted/presented | 2 | Trainees | Completed |
| | Grant applications prepared | 3 | Clinician-scientists | Submitted |

† Participation varied across activities and was not captured as a single cumulative metric; the same individuals could participate in more than one activity.

**Supplementary Table S11. Summary of missing data by variable domain**

| Domain | PD patients (N=300) | Healthy controls (N=335) | Primary reason for missingness |
|---|---|---|---|
| Demographic, ancestry, and ethnicity variables | 0% | 0% | Complete ascertainment |
| Family history and consanguinity | 0-1·0% | 0% | Complete ascertainment |
| Lifestyle and environmental exposures (core) | 0·3-2·0% | 0-0·9% | Minimal participant non-response |
| Nasway use | 88/300 (29·3%) | 3/335 (0·9%) | Item introduced after study initiation; occasional non-response |
| Comorbidities | 0-0·7% | 0-1·2% | Minimal participant non-response |
| RBD1Q | 0% | 3/335 (0·9%) | Minimal non-response |
| Self-reported hyposmia | 0% | 1/335 (0·3%) | Minimal non-response |
| NMSQuest item-level responses | Item-level missingness generally ≤1·3%, except sexual interest 7·7% and sexual difficulty 11·0% | - | Higher non-response to two sexual-function items, primarily because some participants were reluctant or unable to provide an applicable response. |
| Core PD clinical characteristics | 0-2·7% | - | Mostly complete; occasional visit-level incompleteness (CISI-PD) |
| Cognitive assessment (MoCA) | Total score complete: 172/300 (57·3%); missing: 128/300 (42·7%) | - | Phased implementation; baseline completion was limited by trained assessor availability and visit-time constraints. Additional assessments are being obtained during follow-up and an embedded substudy. |

* Missing data include participant non-response (including “prefer not to answer” or “I don’t know”) and assessments or items not administered. Percentages are based on group-specific denominators and, where ranges are shown, represent the minimum and maximum missingness across variables within that domain. Higher missingness for nasway use and MoCA reflects phased implementation rather than loss of previously collected data. For NMSQuest, unanswered sexual-function items were treated as missing and were not assumed to indicate absence of symptoms. Variables not listed separately had minimal or no missing data.

**Supplementary Table S12. Members of the Kyrgyzstan Parkinson's disease research network and participating institutions**

| Institution | Contributors |
|---|---|
| I. K. Akhunbaev Kyrgyz State Medical Academy, Bishkek, Kyrgyzstan | Cholpon Shambetova, Suium Kiyalbekova, Bermet Nurbekova, Elmira Mamytova, Anara Toktomametova, Aleksandr Sorokin, Altynai Baitelieva, Aruuke Kubanychbekova, Amira Sadyrova, Kseniya Taranenko, Islam Sadyrbekov, Gulmira Djumalieva, Indira Kudaibergenova |
| Academic Clinic No. 7, I. K. Akhunbaev Kyrgyz State Medical Academy, Bishkek, Kyrgyzstan | Eliza Zhunusova, Mirbek Baisalov |
| Osh State University, Osh, Kyrgyzstan | Aizhamal Ismailova, Aiperi Toichieva, Mukhammadiusuf Abdykadyrov |
| Kyrgyz State Technical University named after I. Razzakov, Bishkek, Kyrgyzstan | Adilet Metinov, Nursultan Bolotbek uulu, Timur Turatali, Gulnara Kabaeva |
| Department of Medical Care Organization and Drug Policy, Ministry of Health of the Kyrgyz Republic, Bishkek, Kyrgyzstan | Aida Bolotalieva |
| Medical Center "Unimed Clinic", Bishkek, Kyrgyzstan | Ekaterina Peredereeva, Aiza Upaeva |
| Osh City Clinical Hospital, Osh, Kyrgyzstan | Aisuikum Abdumalikova, Nurbakyt Kadyrov, Adylbek Iusupov, Gulstan Noruzbaeva |
| Clinical Hospital under the Presidential Administration of the Kyrgyz Republic, Bishkek, Kyrgyzstan | Nuraiym Beishembieva, Eliza Zhumadylova, Gulmira Zhamilova, Shohista Bazarbaeva |
| Department of Molecular Genetic Research, AquaLab LLC, Bishkek, Kyrgyzstan | Zhyldyz Imanalieva |
| Neurology Department, Chui Regional Hospital, Bishkek, Kyrgyzstan | Aizhamal Iliazova |
| Medical Center "Vedanta" Clinic, Bishkek, Kyrgyzstan | Zhumagul Osmonova |
| Neurology Department, Medical Center "Umai" Clinic, Jalal-Abad, Kyrgyzstan | Gulnaz Pakhridinova, Gulzavera Mamrazhapova |
| National Hospital of the Ministry of Health of the Kyrgyz Republic, Bishkek, Kyrgyzstan | Aziza Dalbaeva, Gulnara Busurmankulova, Karlygach Mamytova, Altynkul Osmonbekova, Saule Temirbaeva |
| Neurology Department, International School of Medicine, Bishkek, Kyrgyzstan | Aliia Mukhanova |
| Department of Neurology and Neurosurgery, S.B. Daniyarov Kyrgyz State Medical Institute of Retraining and Advanced Training, Bishkek, Kyrgyzstan | Irina Sverdlova, Irina Tsopova |
| Neurology Department, Batken Regional United Hospital, Batken, Kyrgyzstan | Batma Sattarova |
| Movement Disorders and Parkinson's Disease Alliance, Bishkek, Kyrgyzstan | Nadira Isakbekova, Aiperi Sydygalieva |
| Neurology Department, Clinical Hospital for Emergency Medical Care, Bishkek, Kyrgyzstan | Nargiza Atambekova |
| Medical Centre "Medcenter.kg", Bishkek, Kyrgyzstan | Sabina Baltabaeva |

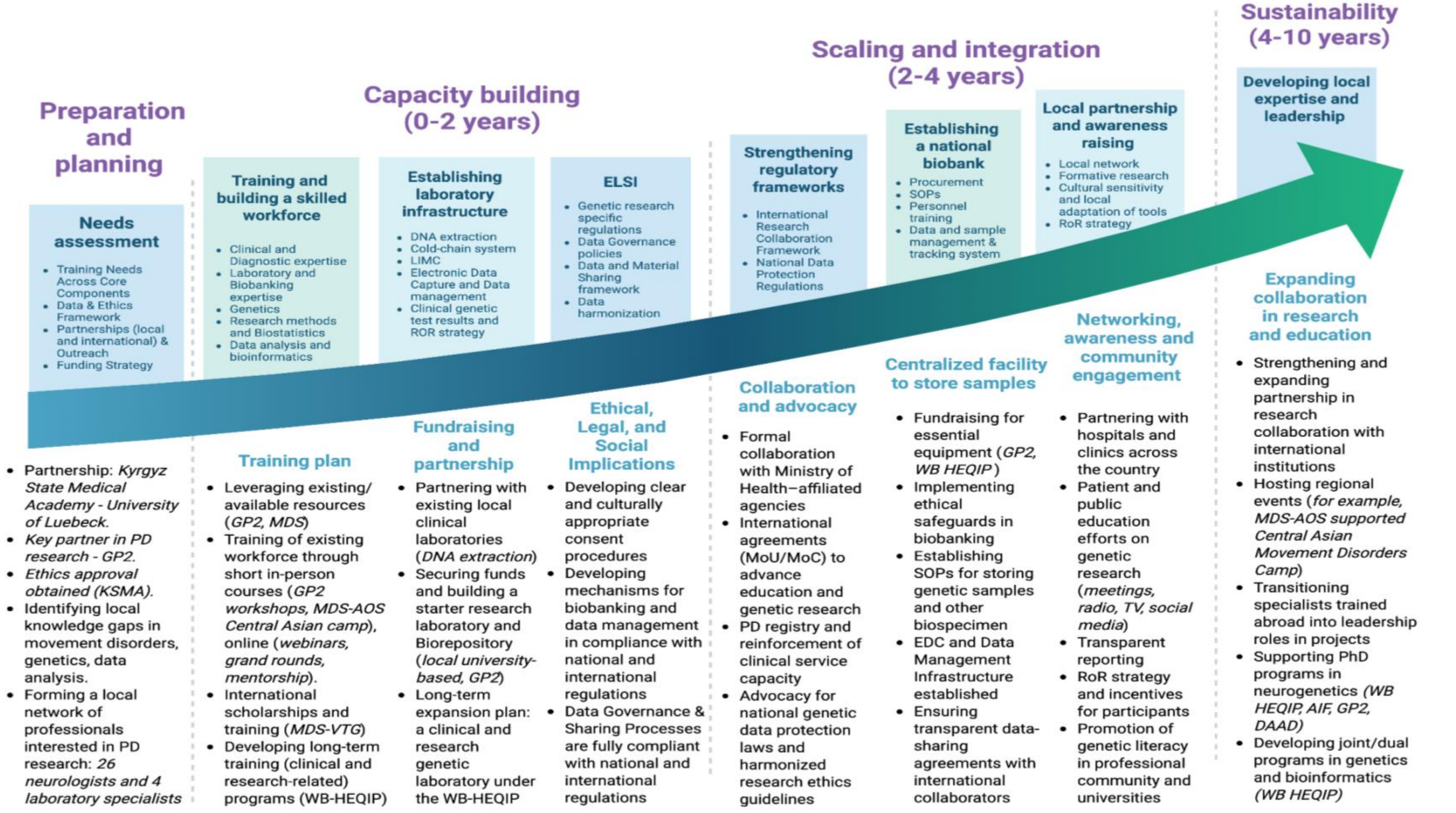

**Supplementary Figure S1. Operational implementation of the framework in Kyrgyzstan.**

The figure provides examples of activities undertaken at each stage of the capacity-building framework in the Kyrgyzstan programme, including training, infrastructure development, governance processes, community engagement, and partnerships. These examples are context-specific and are intended to demonstrate how the framework can be operationalised in practice, rather than to define a required sequence or set of activities.

Created with https://BioRender.com
